# Prevalence of SARS-CoV-2 Antibodies from a one-year National Serosurveillance of Kenyan Blood Transfusion Donors

**DOI:** 10.1101/2021.07.06.21260038

**Authors:** Sophie Uyoga, Ifedayo M.O. Adetifa, Mark Otiende, John N. Gitonga, Daisy Mugo, James Nyagwange, Henry K. Karanja, James Tuju, Johnstone Makale, Rashid Aman, Mercy Mwangangi, Patrick Amoth, Kadondi Kasera, Wangari Ng’ang’a, Nduku Kilonzo, Evelynn Chege, Christine Yegon, Elizabeth Odhiambo, Thomas Rotich, Irene Orgut, Sammy Kihara, Christian Bottomley, Eunice W. Kagucia, Katherine E. Gallagher, Anthony Etyang, Shirine Voller, Teresa Lambe, Daniel Wright, Edwine Barasa, Benjamin Tsofa, Joseph Mwangangi, Philip Bejon, Lynette I. Ochola-Oyier, George M. Warimwe, Ambrose Agweyu, J. Anthony G. Scott

## Abstract

In tropical Africa, SARS-CoV-2 epidemiology is poorly described because of lack of access to testing and weak surveillance systems. Since April 2020, we followed SARS-CoV-2 seroprevalence in plasma samples across the Kenya National Blood Transfusion Service. We developed an IgG ELISA against full length spike protein. Validated in locally-observed, PCR-positive COVID-19 cases and in pre-pandemic sera, sensitivity was 92.7% and specificity was 99.0%. Using sera from 9,922 donors, we estimated national seroprevalence of SARS-CoV-2 antibodies at 4.3% in April-June 2020 and 9.1% in August-September 2020. Kenya’s second COVID-19 wave peaked in November 2020. Here we estimate national seroprevalence in early 2021.

Between January 3 and March 15, 2021, we collected 3,062 samples from donors aged 16-64 years. Among 3,018 samples that met our study criteria, 1,333 were seropositive (crude seroprevalence 44.2%, 95% CI 42.4-46.0%). After Bayesian test-performance adjustment and population weighting to represent the national population distribution, the national estimate of seroprevalence was 48.5% (95% CI 45.2-52.1%). Seroprevalence varied little by age or sex but was higher in Nairobi (61.8%), the capital city, and lower in two rural regions.

Almost half of Kenya’s adult donors had evidence of past SARS-CoV-2 infection by March 2021. Although high, the estimate is corroborated by other population-specific estimates in country. Between March and June, 2% of the population were vaccinated against COVID-19 and the country experienced a third epidemic wave. Natural infection is outpacing vaccine delivery substantially in Africa, and this reality needs to be considered as objectives of the vaccine programme are set.

## Introduction

For high-income countries like Israel and the USA, vaccination has provided an ‘exit’ from the COVID-19 pandemic. For example, in surveillance of blood transfusion donors in the UK, 79% of adults had antibodies to SARS-CoV-2 by June 6, 2021 and 15% had serological evidence of natural infection, indicating that most population immunity was vaccine-derived^1^. Global inequity in COVID-19 vaccine distribution was highlighted at the G7 Summit which committed an extra one billion doses to low-income countries. This focus on doses rather than timing overlooks the pace of transmission in these settings.

Since April 2020, we have undertaken surveillance of blood transfusion donors (aged 16-64 years) in Kenya^2,3^. Using sera from 9,922 donors, national seroprevalence of SARS-CoV-2 antibodies was estimated at 4.3% in April-June 2020^3^, and 9.1% in August-September 2020^2^. Here we estimate seroprevalence for January-March 2021.

## Methods

Plasma samples were collected from all 6 regional transfusion centres and assayed for anti-spike IgG using ELISA^3^. Validated among 910 pre-pandemic sera from coastal Kenya and 174 PCR-positive patients from Nairobi, specificity was 99.0% and sensitivity was 92.7%. We used Bayesian Multi-level Regression with Post-stratification (MRP) to adjust for the age, sex and regional distribution of blood donors compared to national figures. Donors were specified at county level but analysed in regions (adjacent counties)^3^. The surveillance was approved by the Scientific and Ethics Review Unit of Kenya Medical Research Institute (Protocol SSC 3426).

## Results

Between January 3 and March 15, 2021, we collected 3,062 samples (median sample date February 14). Sample numbers from each regional transfusion centre were 1145 (Nairobi), 879 (Mombasa), 431 (Kisumu), 250 (Embu), 200 (Nakuru) and 157 (Eldoret). Forty-four samples were excluded for reasons including missing information, age-ineligible donors, and collection in 2020 (see Figure). Of 3,018 remaining samples, 1,333 were seropositive giving a crude seroprevalence of 44.2% (95% CI 42.4-46.0, Table).

**Figure.**
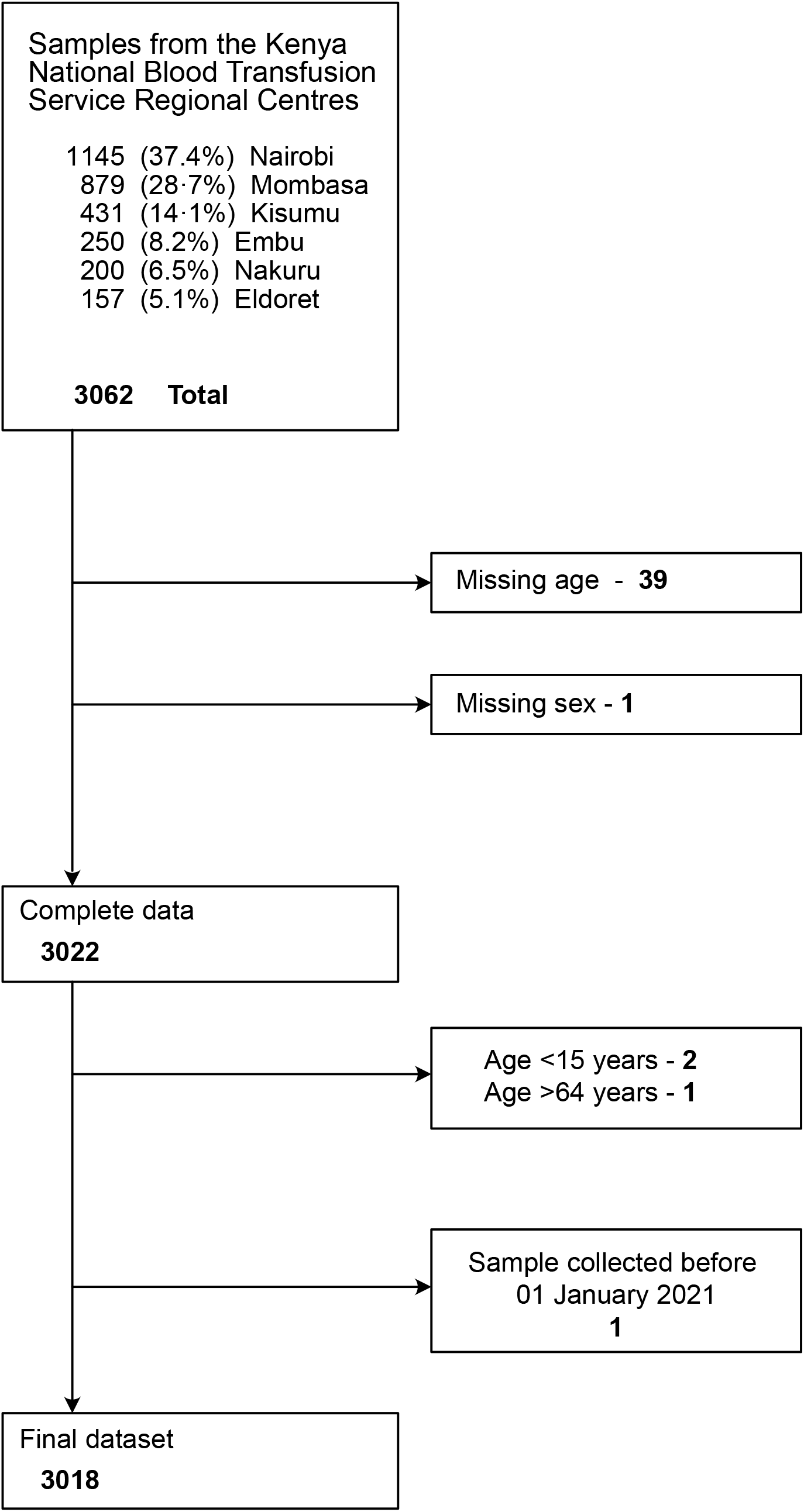
Participant recruitment and flow diagram illustrating exclusions and incomplete data.

**Table.**
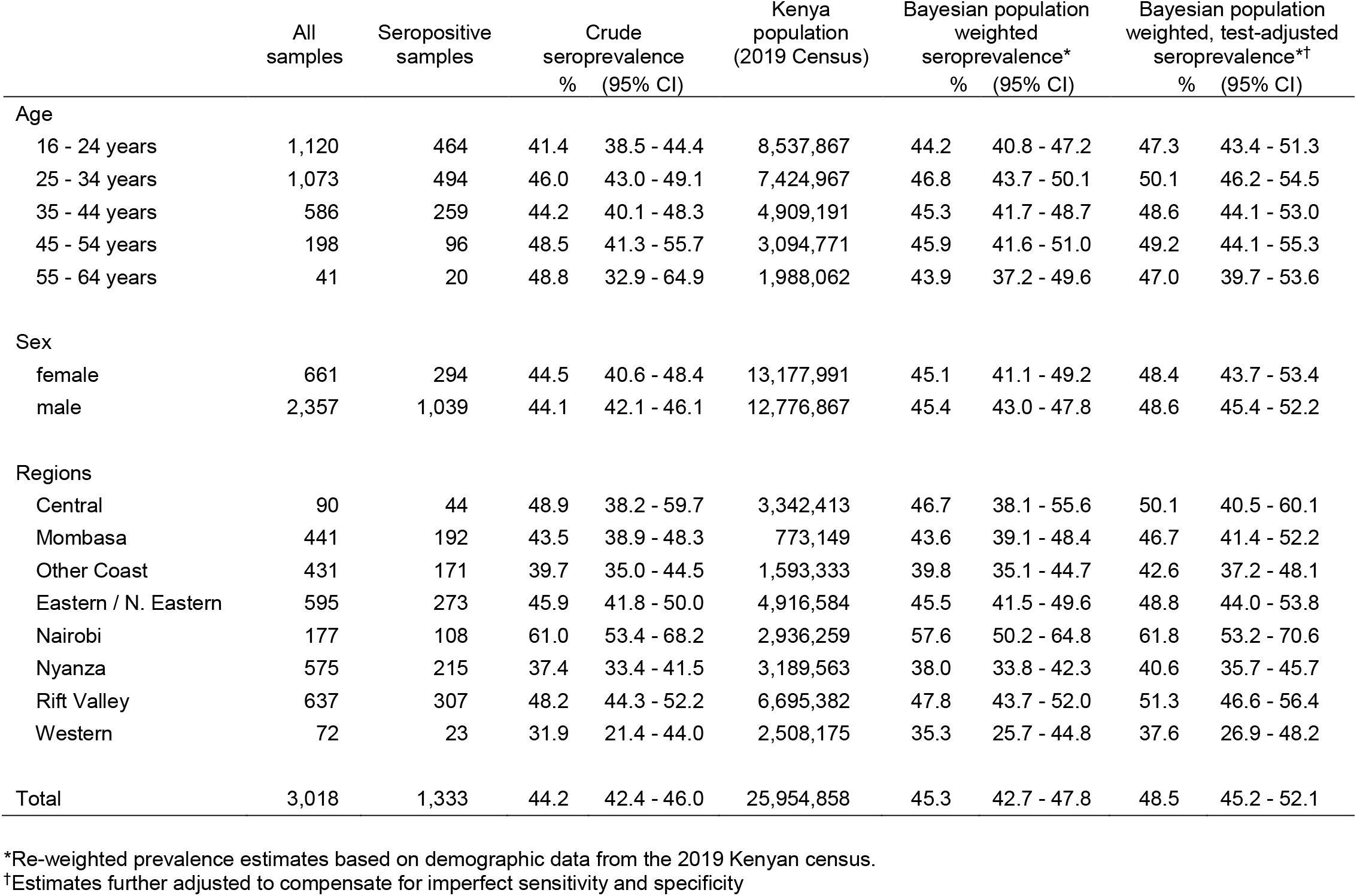
Crude and Bayesian population weighted seroprevalence of anti-SARS-CoV-2 IgG among blood transfusion donors in Kenya by age, sex and region.

After Bayesian population weighting and test-performance adjustment, the estimate of seroprevalence among adults 16-64 years in Kenya was 48.5% (95% CI 45.2-52.1%). This estimate varied little by age or sex but was higher in Nairobi, the country’s capital city and lower in two rural regions in western Kenya, adjacent to Uganda.

## Discussion

Blood transfusion donors are a convenience sample and may not represent the population as a whole, though they have provided useful epidemic intelligence in high income countries^1^. In Kenya, they provide an estimate of cumulative incidence which has grown from 4.3% to 9.1% to 48.5% over a period of 12 months^2,3^. This is consistent with estimates in other populations in Kenya: 35% among Nairobi residents sampled randomly in November 2020; 11% and 50% among antenatal clinic attendees in rural Kilifi and urban Nairobi, respectively, in August-September 2020; 42% among truckers in August-November 2020^2,4^.

The antibody assay was highly specific in Kenyan pre-pandemic samples and demonstrated consistent discrimination in a WHO multi-laboratory standardization exercise^5^. Sensitivity was estimated in individuals sampled a median 21 (minimum 7) days after a positive PCR-test. Antibody concentration, and test sensitivity, may decline with longer times after infection, implying that seroprevalence may be underestimated by our test-performance adjustments.

Collectively these data suggest SARS-CoV-2 has progressed rapidly across Kenya and that half of all adults (16-64 years) were infected by February 2021, before a large third wave of SARS-CoV-2 infections began in March 2021. Kenya’s COVID-19 vaccine programme also began in March 2021 and has reached 2% of the population to date. Elsewhere in Africa, data are sparse but high seroprevalence has been demonstrated; for example, 38% in Juba, South Sudan in October 2020^6^. Natural infection is outpacing vaccine delivery in Africa, and this reality needs to be considered as objectives of the vaccine programme are set.

## Data Availability

The data shown in the manuscript are available upon request from the corresponding author. De-identified data has been published on the Havard dataverse server

https://doi.org/10.7910/DVN/3E1YSQ

## Author Contributions

All authors had full access to all the data in the study and take responsibility for the integrity of the data and the accuracy of the data analysis.

Concept and design: All authors.

Acquisition, analysis, or interpretation of data: All authors.

Drafting of the manuscript: Otiende, Scott. Critical revision of the manuscript for important intellectual content: All authors

Statistical analysis: Otiende.

Obtained funding: Agweyu, Scott, Adetifa, Warimwe.

Administrative, technical, or material support: Uyoga, Adetifa, Warimwe, Yegon, Aman

## Conflicts of Interest

Rashid Aman, Mercy Mwangangi, Patrick Amoth, Kadondi Kasera are from the Ministry of Health in Kenya. Wangari Ng’ang’a is from the Presidential Policy & Strategy Unit of the Government of Kenya. All other authors declare no competing interests.

## Funding/Support

This project was funded by the Wellcome Trust (grant numbers 220991/Z/20/Z, 203077/Z/16/Z), the Bill and Melinda Gates Foundation (INV-017547) and by the Foreign Commonwealth and Development Office (FCDO) through the East Africa Research Fund (EARF/ITT/039). Ambrose Agweyu is funded by a DFID/MRC/NIHR/Wellcome Trust Joint Global Health Trials Award (MR/R006083/1), J. Anthony G. Scott is funded by a Wellcome Trust Senior Research Fellowship (214320) and the NIHR Health Protection Research Unit in Immunisation, Ifedayo Adetifa is funded by the UK Medical Research Council through an African Research Leader Fellowship (MR/S005293/1) and by the NIHR-MPRU at UCL (grant 2268427 LSHTM). GMW is supported by a fellowship from the Oak Foundation. For the purpose of Open Access, the author has applied a CC-BY public copyright licence to any author accepted manuscript version arising from this submission. This paper has been published with the permission of the Director, Kenya Medical Research Institute. We thank all the blood donors for their contribution to the research.

## Data availability

The data shown in the manuscript are available upon request from the corresponding author. De-identified data has been published on the Havard dataverse server https://doi.org/10.7910/DVN/3E1YSQ

## Notes

### Competing Interest Statement

Rashid Aman, Mercy Mwangangi, Patrick Amoth, Kadondi Kasera are from the Ministry of Health in Kenya. Wangari Nganga is from the Presidential Policy and Strategy Unit of the Government of Kenya. All other authors declare no competing interests.

### Author Declarations

The surveillance was approved by the Scientific and Ethics Review Unit of Kenya Medical Research Institute (Protocol SSC 3426).

